# A machine learning model to support the screening for methods guidance articles in MEDLINE: A performance evaluation of ASReview simulation mode

**DOI:** 10.1101/2025.10.13.25337935

**Authors:** Wael Abdelkader, Daniel Xie, Cynthia Lokker, Lingyang Chu, Stefan Schandelmaier, Ashirbani Saha, Muhammad Afzal, Alfonso Iorio

## Abstract

**Background:** Advances in clinical research methods are frequently published in biomedical journals, but identifying these articles remains challenging due to their rapid growth and insufficient indexing in biomedical databases. These challenges hinder the curation of methodologically focused resources like the Library of Guidance for Health Scientists (LIGHTS). Traditional screening approaches, such as Boolean search strategies and manual abstract screening, are inefficient and resource-intensive, limiting the feasibility of regularly updating LIGHTS. Machine learning (ML), particularly active learning models, presents a promising solution to improve the efficiency of article screening.

**Objectives:** This study evaluates the performance of ASReview’s active learning feature in identifying relevant methods guidance articles using pre-labeled data in simulation mode.

**Methods:** Using pre-labeled dataset composed of 1500 methods guidance articles and 20000 clinical studies, categorized as relevant or irrelevant, we trained and compared multiple simulation models in ASReview using various classifiers and feature extraction models. These included combinations of Support Vector Machine (SVM), Naïve Bayes (NB), Neural Network with Sentence BERT (sBERT), Doc2Vec, and TF-IDF. Model performance was evaluated based on screening burden, recall, Work Saved over Sampling (WSS), and precision. All model combinations used maximum query and dynamic double sampling settings.

**Results:** At 95-99.5% recall, SVM with TF-IDF required the fewest screened records (6.87-7.66% burden), while SVM with Doc2Vec achieved the best overall performance at 100% recall with only 11.47% screening burden (WSS@100 = 88.5%) in 42 minutes. Models using sBERT for feature extraction performed comparably through 99.5% recall but exhibited severe performance degradation at 100% recall, requiring screening of over 65% of the corpus.

**Conclusion:** Classical feature extraction methods, TF-IDF and Doc2Vec, paired with SVM outperform deep learning embeddings methods. ASReview in this controlled setting is a feasible tool for screening methodological literature. Future work should include prospective, human-in-the-loop experiments that embed the Doc2Vec-based SVM pipeline in comparison to human screening.

## Introduction

Methods guidance articles are a primary resource for health researchers and are essential to minimize research inefficiencies and avoid research waste. These publications offer advice on designing, conducting, analyzing, interpreting, reporting, or assessing health research studies ^1^. Leading methodologists regularly publish methods guidance articles in top medical journals but also in specialized clinical and methodological journals. Identifying these articles in general biomedical databases such as MEDLINE can be very challenging due to the lack of validated search filters to separate methodological from non-methodological articles, insufficient indexing terms and indexing of methods papers, and highly variable reporting standards ^2^. This is exacerbated by the exponential growth in published biomedical literature at a rate of at least one new article every 26 seconds ^3^. In 2022, over 1.3 million new citations were indexed in MEDLINE, almost double the citations added 5 years prior ^4^.

The Library of Guidance for Health Scientists (LIGHTS, www.lights.science) is a new living online repository of methods guidance articles designed to improve access to methods focused articles for health researchers ^2^. LIGHTS is currently maintained through regular manual screening of selected journals, journal methods sections, and series in selected journals, as well as selected social media accounts and recommendations from researchers which are limited sources for methods guidance articles. A formal Boolean search strategy combined with human screening was piloted and proved not to be feasible due to the very large number of records to be screened^2^. To improve the comprehensiveness and efficiency of updating LIGHTS, a feasible screening procedure is needed.

ML falls within the realm of artificial intelligence ^5^. It is defined as a range of mathematical techniques designed to autonomously recognize patterns in data and subsequently use these patterns to predict future data or other relevant outcomes ^6,7^. In healthcare, machine learning models can effectively identify patterns and relationships, leading to accurate predictions, e.g., patient outcomes and diagnosis ^8,9^. In the biomedical literature field, a primary use of machine learning is text mining—a computer-driven process that extracts novel insights from textual data ^10,11^. Example model functions include classifying studies and recognizing high-quality evidence ^12^, and easing the burden of systematic evidence synthesis by reducing screening workload ^13–16^.

Active learning, also known as "query learning," is a subset of machine learning with the central premise of permitting the learning algorithm to actively choose the data it learns from, displaying a sort of curiosity, to achieve superior performance with a reduced amount of training data ^17^. Active learning is a specialized form of supervised machine learning that allows an algorithm to interactively query a user or another information source to label new data points with desired outputs ^17,18^. Instead of requiring labels for every instance upfront, it begins with a small, labeled dataset and actively selects additional data points to be labeled ^17,19,20^. This approach aims to construct a high-performance classifier while minimizing the training dataset size. The typical process involves starting with a small, labeled dataset, training an initial model, estimating prediction uncertainty, selecting instances with low confidence using various query strategies, obtaining labels from an oracle, updating the model with newly annotated data, and repeating the steps until a performance threshold is reached ^17,18,20–22^. The trained model when applied to unseen data ranks the instances based on their relevance, prioritizing those that are most relevant ^17^. In the context of biomedical literature, the ranking by the model potentially decreases the screening workload by allowing the quicker identification of all relevant documents early, without the need to manually review the complete dataset ^17,20^. Certain groups that maintain living databases for specific methodological areas, such as recruitment methods (Online Resource for Recruitment research in Clinical triAls, ORRCA) and core outcome sets (Core Outcome Measures in Effectiveness Trials, COMET), have successfully implemented learn-to-rank models to enhance their screening processes ^23–25^. Whether such models perform well in a more broadly defined methodological context such as “methods guidance” remains unknown.

## Objective

Using ASReview in simulation mode, our objective is to evaluate the ability of the active learning platform, ASReview, to identify methods guidance articles in MEDLINE. We aimed to compare multiple models’ configurations in terms of screening burden, precision, WSS, and elapsed time at recall levels up to 100 % to identify the most efficient model.

## Methods

### ASReview

The ASReview platform version 1.6.6 was used for developing the active screening models ^20^. ASReview is an open-source software that leverages active learning for model training to enhance the systematic literature review screening process. In this context, active learning involves a ML model working in conjunction with a human reviewer to iteratively select the most pertinent documents from a vast corpus of unreviewed literature ^20^. ASReview platform encompasses various ML classifiers such as logistic regression, Naïve Bayes (NB), Support Vector Machine (SVM) and Convolutional Neural Network (CNN); and feature extraction methods, such as Sentence Bidirectional Encoder Representations from Transformers (sBERT), word embedding, Term-Frequency Inverse Document Frequency (TF/IDF), and Doc2Vec. Using the provided training set, these algorithms analyze patterns in the labeled articles, gaining insights into the characteristics that make an article relevant to the research topic ^20^.

To train active learning screening models, we usually require a small set of labelled articles, which provides an initial training seed for the model and can be as minimal as one relevant and one irrelevant article ^20^. Following this, the ML model selects unlabelled articles with the least certainty and presents them to a human reviewer for assessment. In this iterative process, the human reviewer labels these articles as relevant or irrelevant. For this study, we are utilizing ASReview in simulation mode, in which ASReview’s replays the entire active-learning workflow on a fully labelled corpus. First, the fully labelled dataset is loaded with a small “prior knowledge” seed. Typically, a handful of relevant and irrelevant articles are supplied to initialize the model. The chosen classifier and feature extraction methods are then trained on this seed; the model ranks the remaining unseen records by predicted relevance. Rather than awaiting human judgment for each instance, ASReview immediately consults the concealed pre-labelled data, updates the model with that information, and iterates this loop at machine speed simulating human screening.

Throughout the run ASReview logs the order in which each record is screened, the cumulative recall after every iteration, and the number of records examined, allowing for recall, burden, and WSS calculations at different thresholds. Simulation mode produces perfectly reproducible performance traces for any classifier-feature combination, enabling rapid benchmarking before prospective deployment on unlabeled literature and human screening ^20^.

### Study design

We conducted a simulation-based evaluation of 6 ASReview classifier–feature combinations. Performance was recorded at 95%, 98%, 98.5%, 99%, 99.5%, and 100% recall thresholds. Active-screening performance metrics were reported, including WSS, precision, Burden, and time to achieve 100% recall ^26,27^ (supplementary Table 1). We used 6 different configurations for the models: NB and TF/IDF, SVM and TF/IDF, SVM and Doc2Vec, SVM and sBERT, CNN and Doc2Vec, and CNN and sBERT. We used ten articles as prior knowledge, of which five were relevant and five were irrelevant. The articles used as prior knowledge were fixed across all models and were selected by a random number generator (supplementary Table 3). We used the maximum query strategy combined with a double dynamic resampling balancing strategy for all experiments. All experiments were conducted on a single machine with a 13-generation core i9-13900H CPU, 32Gb of RAM, and Nvidia RTX-4070 GPU.

### Dataset description

We used a pre-labelled dataset consisting of 21,535 PubMed articles, of which 1,535 articles were retrieved from the LIGHTS database, and they represented the “relevant” class ^2^. Articles are included in the LIGHTS database if they meet the following inclusion criteria at full-text screening:

1. Published in a peer-reviewed journal.
2. Title, series title, section title, author keywords, or abstract states aim to provide guidance on a methodological topic. Different terms for guidance are accepted, such as tutorial, recommendation, best practice, among others.
3. The articles address methods relevant to health-related research involving humans.

The remaining 20,000 articles representing the “irrelevant” class were retrieved from the McMaster PLUS database which excludes methods guidance articles as one of its inclusion criteria. The McMaster PLUS dataset includes titles and abstracts of 155,679 articles published between 2012 and 2024. These articles were identified using PubMed identifiers developed by the Health Research Information Unit (HiRU) at McMaster University and were labeled by expert research associates ^28,29^.

## Results

The number of articles screened by the different classifier feature combinations at the various recall thresholds for relevant articles (95%, 98%, 98.5%, 99%, 99.5%, and 100%) are shown in Table 1.

**Table 1:** Screening performance for models tested configurations at defined recall thresholds in the ASReview simulation.

| Classifier | Feature extraction | Total records screened | Time to achieve 100% recall |
| --- | --- | --- | --- |

|  |  | **95%**<br>**recall**<br>**(n=**<br>**1458)** | **98%**<br>**recall**<br>**(n=**<br>**1504)** | **98.5%**<br>**recall**<br>**(n=**<br>**1512)** | **99%**<br>**recall**<br>**(n=**<br>**1520)** | **99.5%**<br>**recall**<br>**(n=15**<br>**27)** | **100%**<br>**recall**<br>**(n=15**<br>**35)** |  |
| --- | --- | --- | --- | --- | --- | --- | --- | --- |
| **NB** | TF/IDF | 1650 | 1990 | 2120 | 2480 | 3160 | **3930** | 1 minute |
| **SVM** | TF/IDF | <u>1480</u> | 1560 | 1590 | 1620 | 1650 | **2900** | 15 hour and 31 minutes |
| **SVM** | Doc2Vec | 1540 | 1690 | 1730 | 1790 | 1920 | **2470** | 42 minutes |
| **SVM** | sBERT | 1500 | 1580 | 1610 | 1660 | 1690 | **14460** | 11 hours and 5 minutes |
| **NN** | Doc2Vec | 1550 | 1670 | 1790 | 1990 | 2420 | **7240** | 33 hours and 6 minutes |
| **NN** | sBERT | 1510 | 1570 | 1590 | 1630 | 1820 | **13950** | 45 hours and 14 minutes |

At the 95% recall threshold, the SVM paired with TF-IDF required the fewest screened records (n=1480). The next best configurations were sBERT combined with SVM classifier and CNN with each requiring 1500 and 1510 records, respectively. These three models remained the top three performing models till the 99.5% recall threshold with the SVM+TF/IDF remaining the best performing while the two sBERT models alternated between second and third place separated by only a small margin. At 100% recall, the two SVM-based models with classical feature extraction set the efficiency benchmark with SVM + Doc2Vec reached 100% recall after reviewing only 2470 records, 11.47% burden, yielding a WSS@100 of 88.5%. Precision decreased from 94.7% at 95% recall to almost 62% at 100% recall, however, the SVM+Doc2Vec model completed the task in just 42 minutes. SVM + TF-IDF was the second-best performing mode, with a marginally lower WSS@100 of 86.5% and a longer time of 15 hours 31 minutes; nevertheless, its precision remained the highest below 100 % recall (Table 2).

**Table 2:** Models’ screening burden, precision, and WSS performance metrics at recall thresholds.

| **Model** | **Recall threshold (%)** | **Burden (%)** | **Precision (%)** | **WSS (%)** |
| --- | --- | --- | --- | --- |
| **NB + TF-IDF** | 95 | 7.66 | 88.36 | 91.93 |
| **NB + TF-IDF** | 98 | 9.24 | 75.58 | 90.57 |
| **NB + TF-IDF** | 98.5 | 9.84 | 71.32 | 90.01 |
| **NB + TF-IDF** | 99 | 11.52 | 61.29 | 88.37 |
| **NB + TF-IDF** | 99.5 | 14.67 | 48.32 | 85.25 |
| **NB + TF-IDF** | 100 | 18.25 | 39.06 | 81.75 |
| **SVM + TF-IDF** | 95 | 6.87 | 98.51 | 92.77 |
| **SVM + TF-IDF** | 98 | 7.24 | 96.41 | 92.61 |
| **SVM + TF-IDF** | 98.5 | 7.38 | 95.09 | 92.5 |
| **SVM + TF-IDF** | 99 | 7.52 | 93.83 | 92.4 |
| **SVM + TF-IDF** | 99.5 | 7.66 | 92.55 | 92.3 |
| **SVM + TF-IDF** | 100 | 13.47 | 52.93 | 86.53 |
| **SVM + Doc2Vec** | 95 | 7.15 | 94.68 | 92.47 |
| **SVM + Doc2Vec** | 98 | 7.85 | 88.99 | 91.99 |
| **SVM + Doc2Vec** | 98.5 | 8.03 | 87.4 | 91.84 |
| **SVM + Doc2Vec** | 99 | 8.31 | 84.92 | 91.6 |
| **SVM + Doc2Vec** | 99.5 | 8.92 | 79.53 | 91.04 |
| **SVM + Doc2Vec** | 100 | 11.47 | 62.15 | 88.53 |
| **SVM + sBERT** | 95 | 6.97 | 97.2 | 92.67 |
| **SVM + sBERT** | 98 | 7.34 | 95.19 | 92.51 |
| **SVM + sBERT** | 98.5 | 7.48 | 93.91 | 92.41 |
| **SVM + sBERT** | 99 | 7.71 | 91.57 | 92.21 |
| **SVM + sBERT** | 99.5 | 7.85 | 90.36 | 92.11 |
| **SVM + sBERT** | 100 | 67.15 | 10.62 | 32.85 |
| **NN + Doc2Vec** | 95 | 7.2 | 94.06 | 92.42 |
| **NN + Doc2Vec** | 98 | 7.75 | 90.06 | 92.09 |
| **NN + Doc2Vec** | 98.5 | 8.31 | 84.47 | 91.56 |
| **NN + Doc2Vec** | 99 | 9.24 | 76.38 | 90.67 |
| **NN + Doc2Vec** | 99.5 | 11.24 | 63.1 | 88.71 |
| **NN + Doc2Vec** | 100 | 33.62 | 21.2 | 66.38 |
| **NN + sBERT** | 95 | 7.01 | 96.56 | 92.62 |
| **NN + sBERT** | 98 | 7.29 | 95.8 | 92.56 |
| **NN + sBERT** | 98.5 | 7.38 | 95.09 | 92.5 |
| **NN + sBERT** | 99 | 7.57 | 93.25 | 92.35 |
| **NN + sBERT** | 99.5 | 8.45 | 83.9 | 91.51 |
| **NN + sBERT** | 100 | 64.78 | 11 | 35.22 |

By contrast, models using sBERT for feature extraction recorded higher burden and runtime once recall surpassed the 99% threshold. SVM + sBERT and NN + sBERT had to screen more than 14,000 and 13,900 records, respectively, representing over 65% of the corpus, driving precision below 35% and WSS@100 to near-random levels, all while consuming 33 to 45 hours of compute time. The Naïve Bayes model with TF-IDF sat between these extremes needing only a one-minute runtime to reach 100% recall at the price of a higher screening burden of 18.3% and reduced WSS of 81.8% (Table 2).

## Discussion

In this study, we used ASReview simulation engine combined with a fully labelled corpus of 1535 methods guidance articles and 20000 non-guidance citations to evaluate the performance of the assisted screening as a component to reduce the screening workload towards the identification of methods guidance articles. Six classifier-feature extraction configurations were tested at recall thresholds ranging from 95% to 100%. At 95% recall, all models had a similar performance with WSS@95 around 92% with less computationally demanding models achieving the highest performance. This indicates that ASReview can effectively reduce the workload of methods guidance articles screening. At higher recall thresholds up to 99.5% recall, performance was still comparable with the SVM models achieving the best workload reduction indicating negligible benefit from deep feature extraction in this range. At 100% recall threshold, deep feature extraction methods tended to underperform in both the amount of workload burden needed to reach 100% of almost 70% of the dataset, with much longer computational demands in comparison to lightweight models like NB and SVM models. In the study done by Ferdinands et al. (2023), they used multiple models in ASReview simulation and concluded that lightweight models can outperform complex computationally demanding models in terms of WSS@95 and in the time elapsed to complete the screening ^30,31^.

While screening tasks conventionally aim for 100% recall, ensuring all relevant articles are identified, this endpoint offers limited insight into the comparative performance of assisted screening methods. The meaningful distinction between approaches, particularly those employing ML assisted screening, emerges in how efficiently they reduce the screening workload and burden while navigating the most uncertain articles during the screening process. This zone of uncertainty is not fixed but rather dataset-dependent; in one review, the most challenging decisions may cluster around 99% recall as in our case, while in another, ambiguity may peak much earlier at 80% recall due to differences in topic complexity, terminology consistency, or any other reason. Therefore, performance evaluation should focus not on the inevitable endpoint of complete recall, but on how effectively different methods handle uncertainty at whatever recall threshold it occurs.

To date, no experiments have applied ML to identify methods guidance articles ^12,32^. Our examination of ASReview offers an approach offering directions for the integration of ML into the LIGHTS database infrastructure, facilitating the otherwise unmanageable screening of results from biomedical database searches. One of our study strengths is the use of a large, expertly labelled reference standard encompassing both a substantial positive class of methods guidance articles in combination of a large pool of 20,000 irrelevant citations, thereby allowing precise estimates of burden, precision and WSS ^31,33^. The use of the simulation mode is advocated for evaluating active learning models before systematic screening ^34^. The use of ASReview’s simulation with a fully labelled corpus eliminates the inter-reviewer variability, thereby enabling the evaluation of multiple models’ configurations. Under identical conditions, a reproducible head-to-head comparison can be achieved by isolating the models’ performance from reviewer variability ^34^. Furthermore, comparing classifier–feature pairs is an important step towards trustworthy screening automation as screening efficiency at a fixed recall depend not only on the classifier and active learning strategy used but also on how documents are represented. Prior studies show that different classifier-feature choices can affect WSS and screening burden at high recall even under the same dataset and labeling strategy (Ferdinands et al., 2023). ASReview offers multiple algorithms and vectorizers but does not prescribe a default best, making empirical head-to-head testing necessary for transparent, reproducible selection in each review (van de Schoot et al., 2021). Modern embedding models such as sBERT for example capture semantics that bag-of-words or TF-IDF representations may miss, plausibly shifting prioritization curves (Reimers & Gurevych, 2019). This comparison provides evidence directly relevant to resource constrained groups considering whether the computational overhead of deep embeddings is justified over less computationally demanding approaches such as TF-IDF or Doc2Vec embedding (Bailly et al., 2022). Finally, the use of fixed seeds and a publicly available open-source platform ensures that our workflow is fully replicable and readily extendable to other biomedical corpora, thereby enhancing both the transparency and generalizability of our findings. The main limitation is the generalizability of our results given the selective dataset combining methods guidance articles with medical studies. The dataset provided a high contrast between relevant and irrelevant whereas the results of real Boolean searches would typically contain a substantial proportion of difficult-to-classify articles such as methods papers that do not satisfy the criteria for “methods guidance.” Another limitation is the size of the dataset used. Although the 21,535 records dataset is sizeable by ML standards, it does not approximate the scale encountered in real-world methods guidance searches. Hirt et al. found that a keyword-based MEDLINE query for methodological guidance returned more than 915,000 citations ^2^. Consequently, the workload reductions observed here are considered an optimistic step towards the incorporation of ML assisted screening within the LIGHTS database workflow. Our findings suggest a practical path towards future adoption into the LIGHTS database screening process which includes three main steps: model selection, validation, and implementation. Using the simulation mode in ASReview in this study allowed us to select the best performing classifier-feature pair to maximize WSS at high recalls while minimizing screening burden and compute time. Following we aim to validate the selected model using a prospective, human-in-the-loop validation on an unlabeled, mixed-topic dataset to confirm that the selected model achieves the simulated WSS, burden reduction, and precision at desired recall.

## Conclusion

Our study results show that combining an SVM with a lightweight feature representation, Doc2Vec, offers the most favourable balance of low screening burden, high WSS@95 to WSS@100, high precision, and minimal elapsed time, whereas deep-embedding architectures confer little advantage in this simulation and can dramatically inflate workload and turnaround. Future work should include a more real-world representative dataset and prospective, human-in-the-loop experiments that embed the Doc2Vec-based SVM pipeline in comparison to human screening. Future experiments should also explore different stopping criteria driven by informed decisions to balance the risk of missing any relevant articles and the costs of continued screening and its burden.

## Data Availability

The data underlying the results presented in the study are available from McMaster PLUS: https://plus.mcmaster.ca/mcmasterplusdb/ LIGHTS database: https://lights.science/

## Supplementary information

### Search strategy for MEDLINE / Ovid

Below is the MEDLINE search strategy that the LIGHTS team developed. Search date: 28/07/2021; Ovid MEDLINE(R) ALL years 2010 to 2020. Number of hits: **876,395**. Because the number of hits is so large, current search updates of LIGHTS are based on a version of this strategy that is limited to the most relevant journals only.

(guideline OR practice guideline OR consensus development conference OR consensus development conference, NIH).pt.

OR Guidelines as Topic/ OR practice guideline/ OR Checklist/ OR exp standards/ OR exp Consensus/ OR exp Consensus Development Conferences as Topic/ OR exp Delphi Technique/ OR Decision Making/

OR (guideline OR guidance OR guide OR guiding OR Tutorial OR Tutorials OR white paper OR Framework OR Checklist OR Checklists OR step-by-step OR Primer OR pitfall OR Pitfalls OR consensus* OR Delphi OR Expert-panel).ti,ab,kf,kw.

OR ((provide OR providing OR provided OR provision OR give OR giving OR gave OR given OR practical) ADJ2 (advice OR recommend* OR tip OR tips)).ti,ab,kf,kw.

OR (Recommend*).ab. /freq=2

OR ((best OR code OR good) ADJ2 (practice OR practices)).ti,ab,kf,kw.

OR (guidelines OR Standard OR Standards OR Recommend* OR Elaboration OR elaborating OR Explanation OR explaining OR extension).ti,kf,kw.

OR (Statement* OR Principle OR Principles OR Principled OR tool OR tools OR Rule OR Rules OR how-to OR critical-question*).ti.

AND (201*.ez,ep,dt. OR 201*.dp. OR 2020.ez,ep,dt. OR 2020.dp.)

**Supplementary Table 1:** Summary of the performance metrics will be used.

| Metric | Definition | Formula |
| --- | --- | --- |
| **Recall**<br>**(sensitivity)** | Recall calculates the proportion of true positive results (correctly identified as relevant) out of all actual positive results (the total number of relevant articles in the dataset). | $Recall = \frac{\text{True positive}}{\text{Total relevant}}$ |
| **Work saved over sampling (WSS)** | The percentage of papers that the reviewers do not have to read because they have been screened out by the classifier <sup>26</sup>. | $WSS@r\% = \left( 1 - \frac{Burden/100}{Recall} \right) * 100$ |
| **Burden** | Express the percentage of the entire dataset that necessitates manual review by a human when using a particular screening approach <sup>27</sup>. | $Burden = \frac{\text{Total screened}}{\text{Total articles}}$ |
| **Precision** | Express the proportion of the screened records that are relevant <sup>27</sup>. | $Precision = \frac{\text{True positive}}{\text{Total screened}}$ |

**Supplementary table 2:**
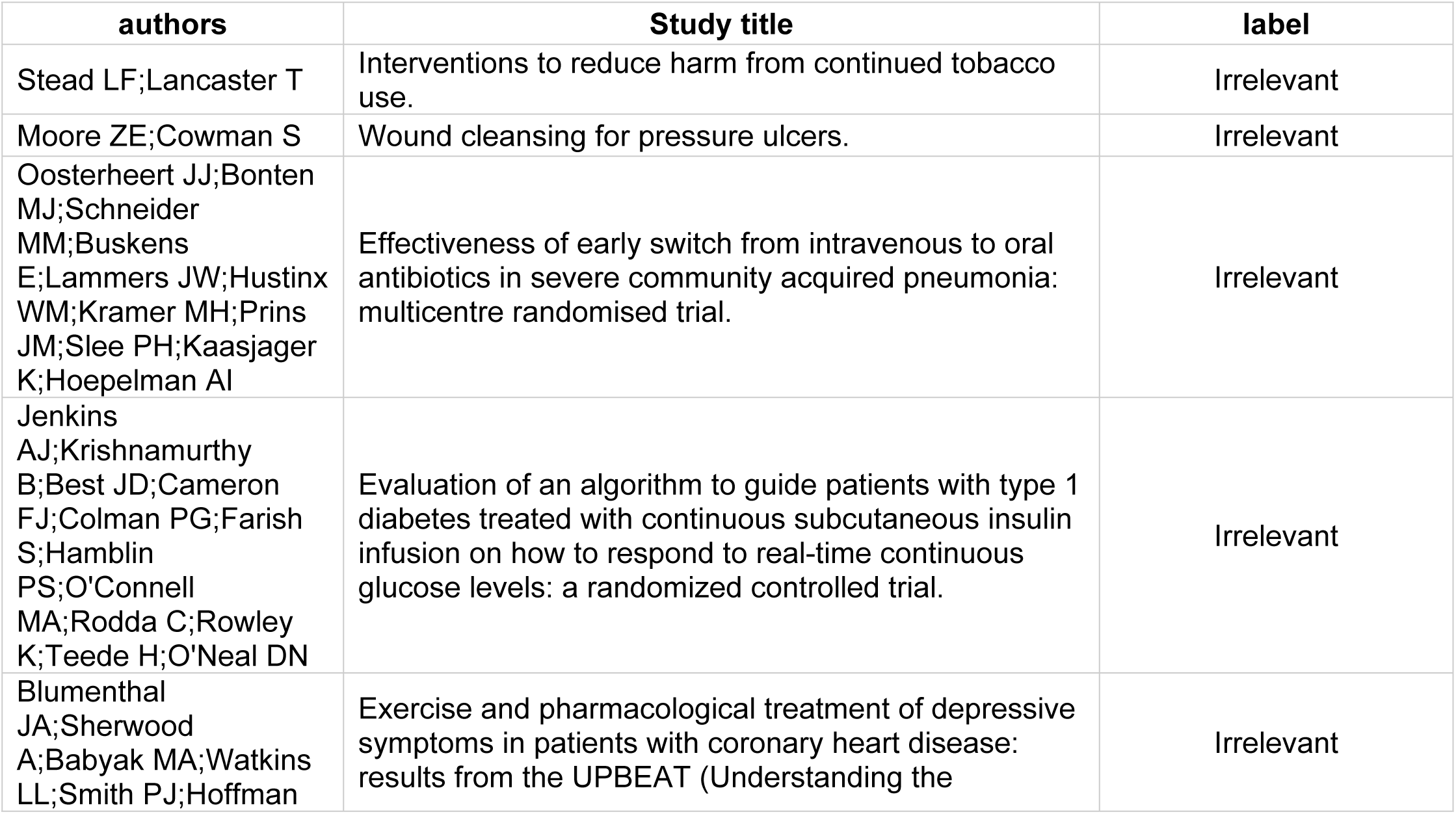
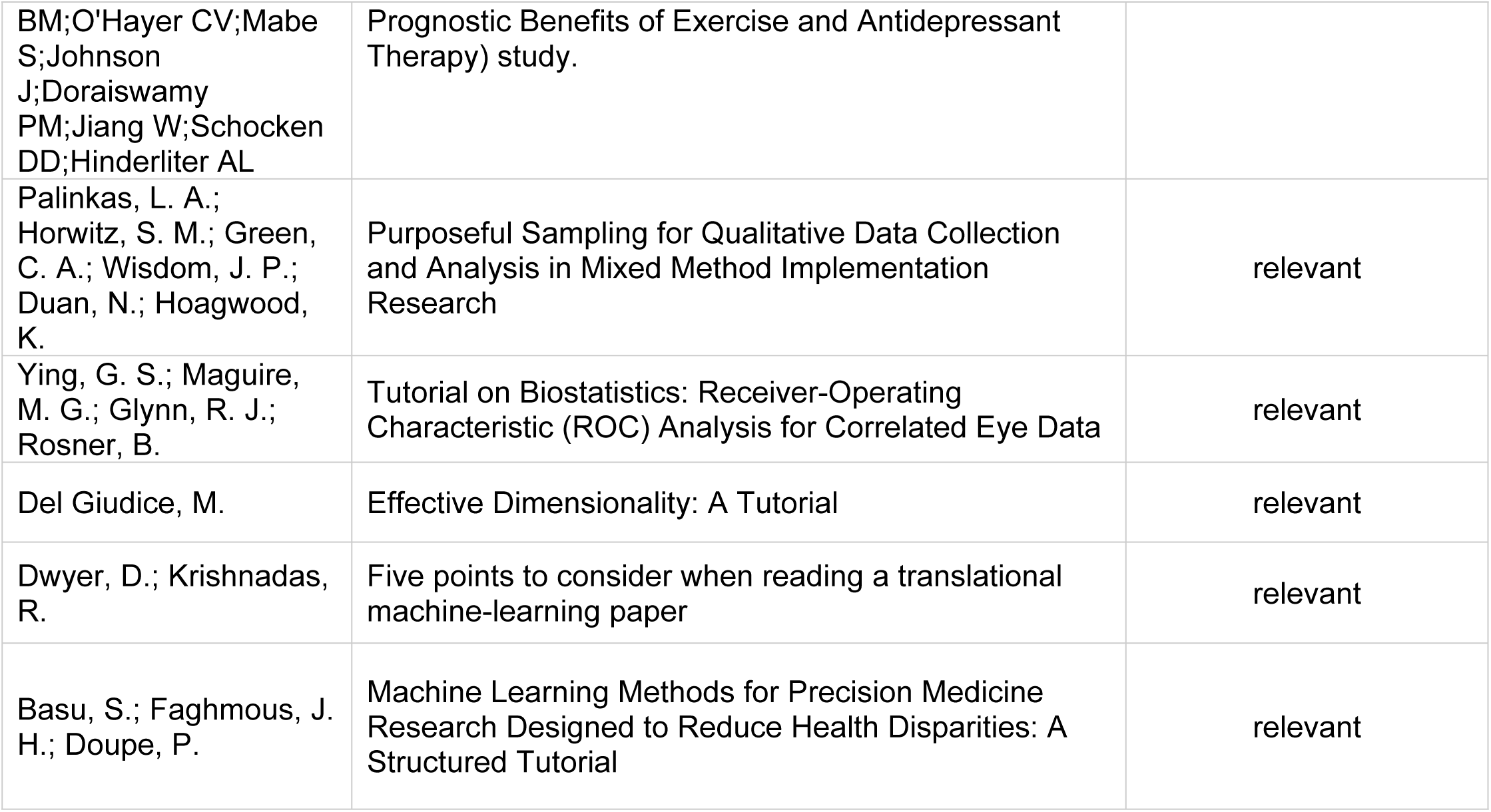
Prior knowledge studies used as fixed seed.

**Supplementary figure 1:**
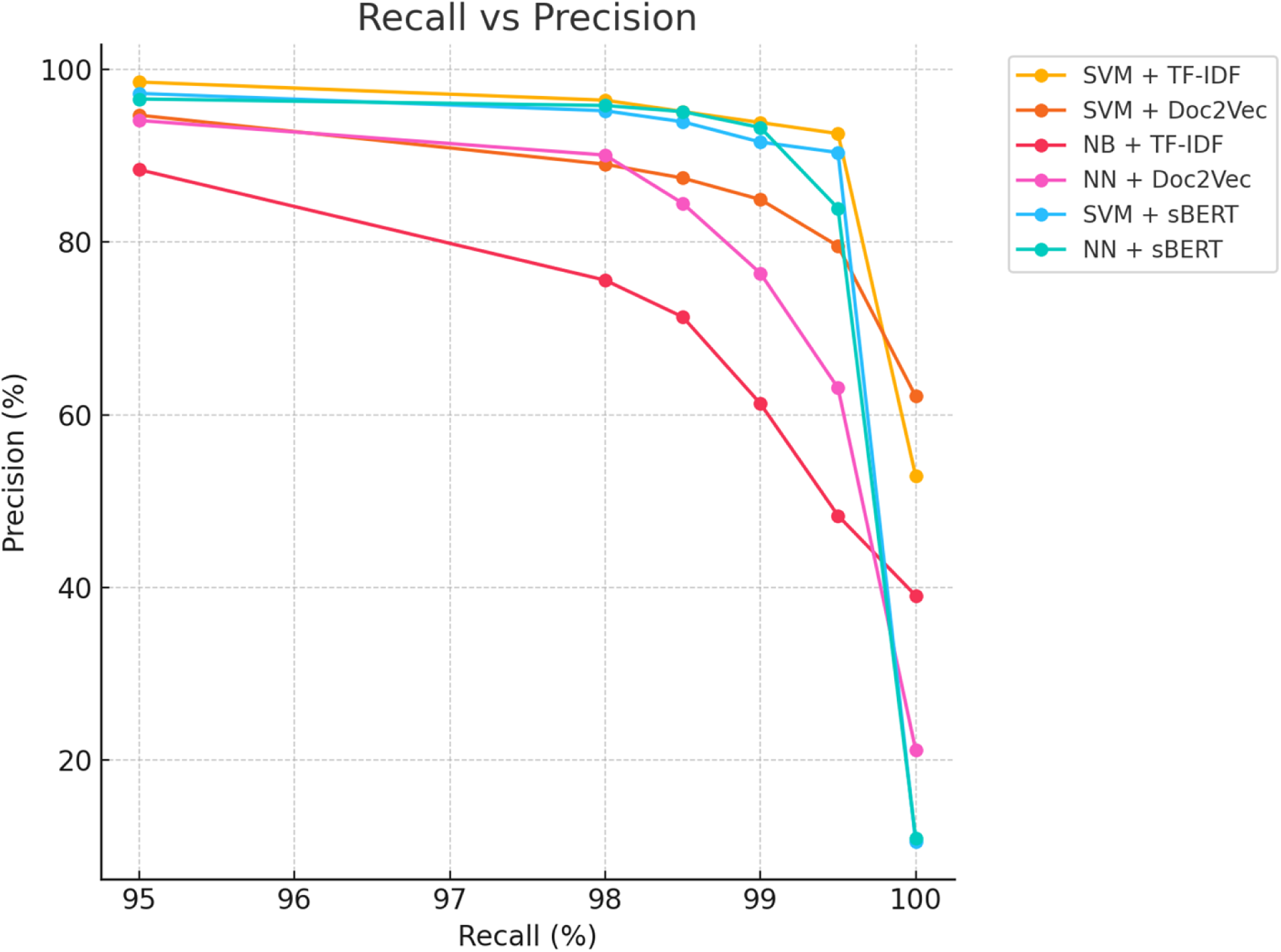
Models’ Precision (%) against Recall (%)

**Supplementary figure 2:**
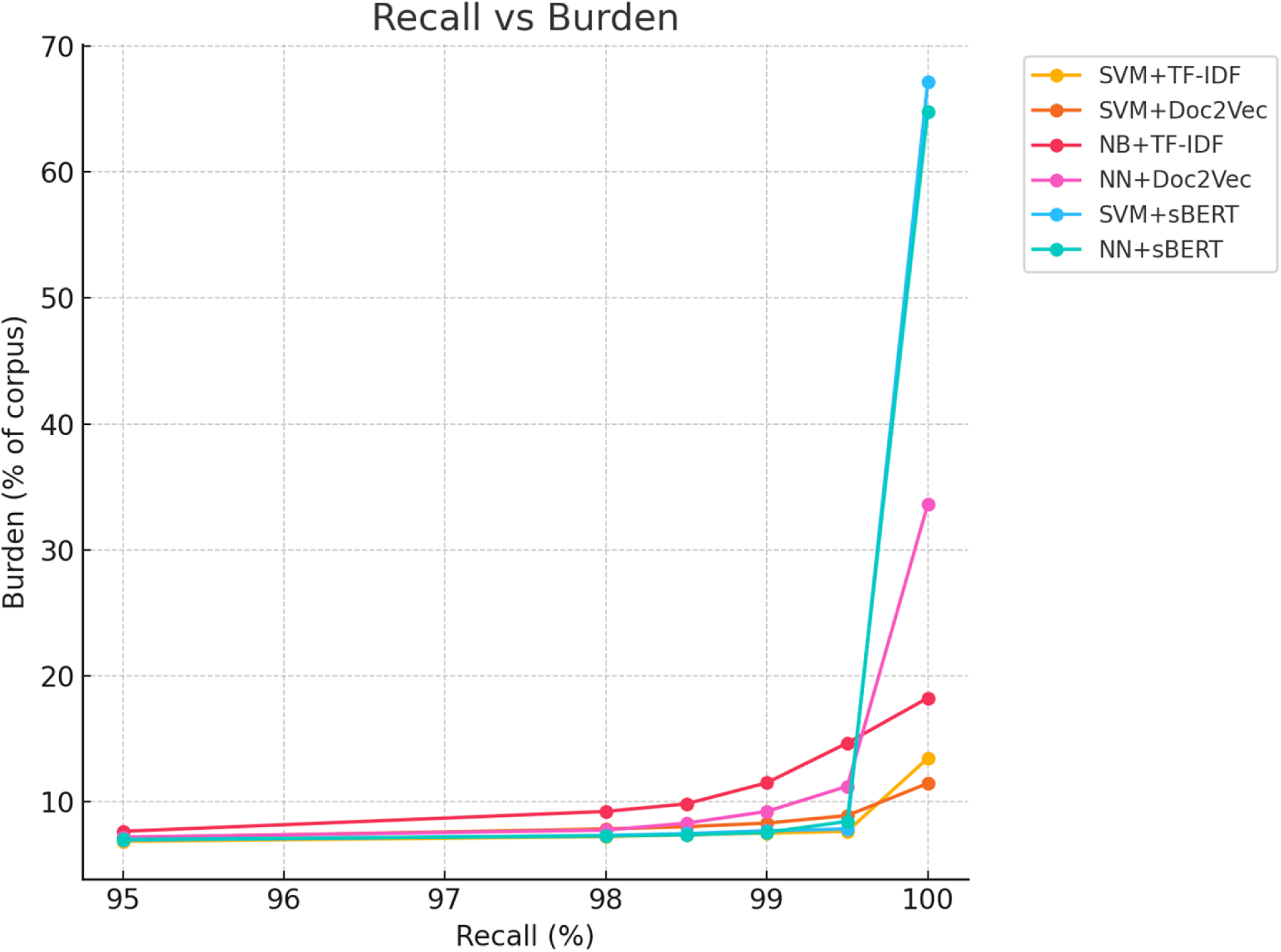
Models’ Burden (%) against Recall (%)

**Supplementary figure 3:**
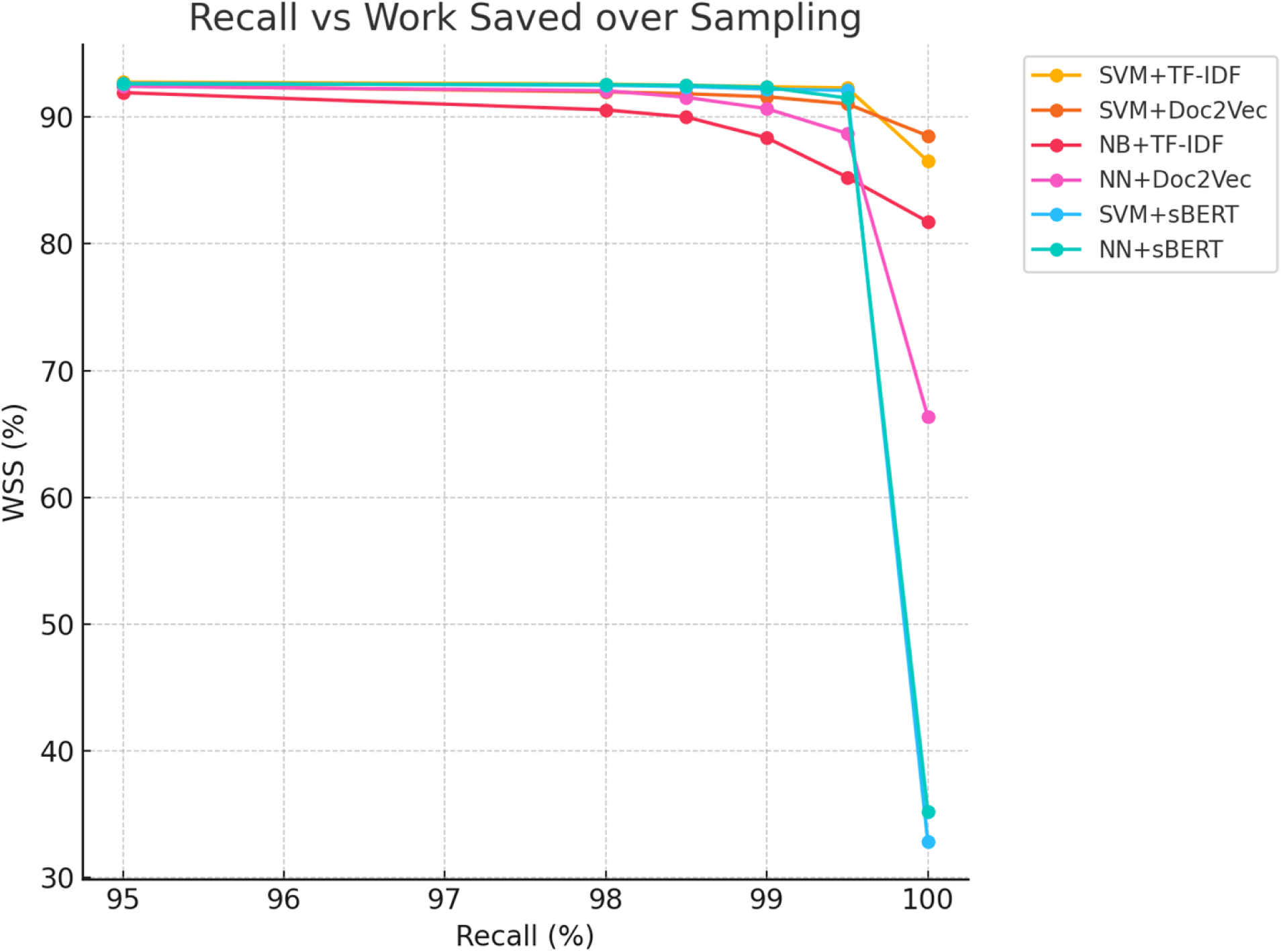
Models’ WSS (%) against Recall (%)

**Supplementary figure 4:**
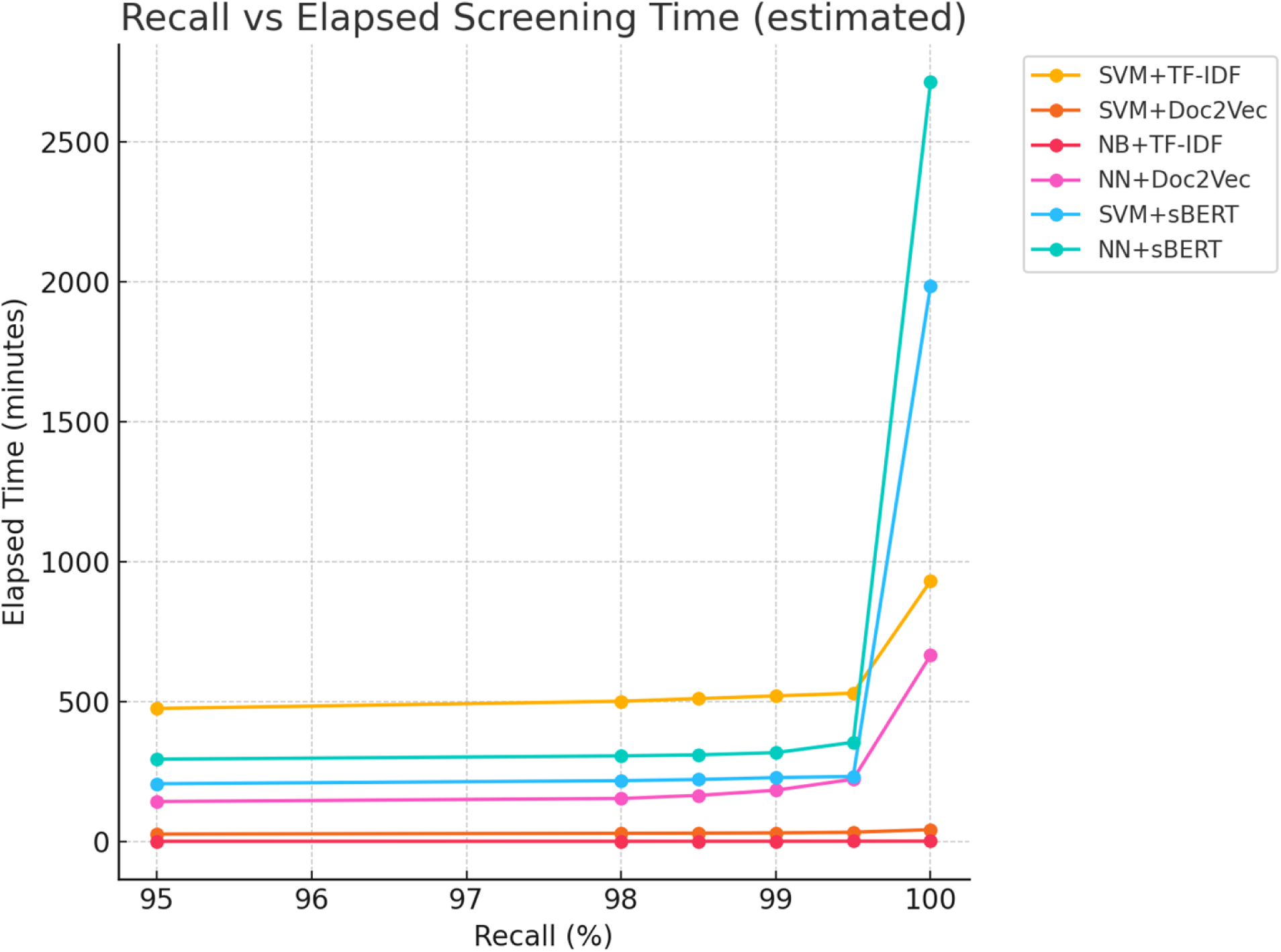
Models’ elapsed screening time (%) against Recall (%)

## Notes

### Competing Interest Statement

The authors have declared no competing interest.

### Funding Statement

The author(s) received no specific funding for this work.

